# Non-COVID-19 Deaths After Social Distancing in Norway

**DOI:** 10.1101/2020.06.05.20123695

**Authors:** Ralph Catalano, Joan A. Casey, Tim A. Bruckner, Alison Gemmill

## Abstract

Lay persons and policy makers have speculated on how the imposition of social distancing to reduce SARS CoV-2 (severe acute respiratory syndrome coronavirus 2) infection has affected non-COVID-19 (coronavirus disease of 2019) deaths. No rigorous estimation of the effect appears in the scholarly literature. We use time-series methods to compare non-COVID-19 deaths observed in Norway before and during the epidemic to those expected from non-COVID-19 deaths in Sweden as well as from the history of Norwegian mortality trends. We find that in the first 6 weeks after the divergence between Swedish and Norwegian policies -- the only period for which dependable data can be had – approximately 414 fewer Norwegians died than expected.

## INTRODUCTION

Intuition and limited data [1] suggest that social distancing intended to reduce SARS-CoV-2 (severe acute respiratory syndrome coronavirus 2) infections may also affect the incidence of death caused by the hazards of “normal” life. Flu infections as well as work and motor vehicle accidents, for example, may decline when social interactions become less common. Such distancing may, however, also increase deaths by reducing social support and observation of frail persons as well as by delaying access to medical care. Formal estimation of the “net effect” of these countervailing mechanisms on the incidence of death would help inform collective choices concerning the management of the pandemic. No such estimates, however, appear in the scholarly literature.

On March 12, 2020, Norway’s government implemented a series of non-pharmaceutical interventions that curtailed social interaction [2]. As widely reported, Sweden’s government chose less restrictive policies [3]. These different approaches taken by neighboring countries with similar socio-economic characteristics provide an opportunity to estimate the reductions in deaths associated with social distancing policies.

We apply time-series methods to compare non-COVID-19 deaths observed in Norway before and during the epidemic to those expected from non-COVID-19 deaths in Sweden as well as from the history of Norwegian mortality trends. We focus, as described below, on the first 8 weeks after the divergence between Swedish and Norwegian policies because more recent data were not available at the time of our analyses.

## METHODS

We obtained weekly counts of deaths for the 538 weeks beginning January 4, 2010 and ending May 3rd, 2020 from the Human Mortality Database’s Short-term Mortality Fluctuations (STMF) data series [4]. As of May 2020, the STMF data contain weekly death counts from 15 countries including Sweden and Norway. Deaths are assigned to weeks based on their date of occurrence and are disaggregated by age and sex. Data from Sweden were provided by Eurostat and Statistics Sweden, while data from Norway were provided from Statistics Norway. The most recent data, updated on June 7, at the time of our analyses stopped with the week ending May 24. Counts for the most recent 3 weeks at each update are considered preliminary. We, therefore, ended our analyses with the week ending May 3, 2020.

We also obtained daily counts of COVID-19 deaths from the Your World in Data Website [5] and aggregated them to weeks. We created our test variables by subtracting COVID-19 weekly deaths from all weekly deaths for both Norway and Sweden.

Our analyses used Box-Jenkins time-series methods [6] adapted for epidemiologic questions [7]. These methods remove from weekly Norwegian deaths the variation predictable not only from deaths in Sweden, but also from autocorrelation (i.e., trends, seasonality, or the tendency to remain elevated or depressed after high or low values) observed in the history of Norwegian deaths before social distancing. This approach rules out confounding by “third variables” affecting both Sweden and Norway as well as those peculiar to Norway that exhibit autocorrelation. The approach also assures a statistically efficient estimation of the association with the imposition of social distancing because the adjusted series will meet the assumption of constant mean and serial independence.

We proceeded through 4 steps, described below, using Box-Jenkins time series modeling.

1. We regressed the weekly number of non-COVID-19 deaths in Norway on those in Sweden for the 538 weeks beginning January 4, 2010 and ending May 3, 2020.
2. We used Box and Jenkins methods to detect autocorrelation including trends, cycles (e.g. seasonality), and/or the tendency to remain temporarily elevated or depressed after high or low values in the residuals of the regression estimated in step 1.
3. We estimated a Box-Jenkins transfer function formed by including parameters specifying autocorrelation detected in step 2 in the model estimated in step 1.
4. We estimated a test equation formed by adding a binary “social distancing” variable to the transfer function model developed in step 3. We scored the binary variable 1 for the week starting March 16th and 0 otherwise. We specified the variable to allow estimating “lagged” effects through 6 following weeks.

## RESULTS

An average of 787 Norwegians, ranging from 663 to 1048, died from non-COVID-19 related causes per week over the 538 weeks of our study. 1717 Swedes on average died from causes other than COVID-19 each week with a range of 1441 to 2204.

Table 1 shows the estimated transfer functions for deaths among Norwegians for the 538 weeks beginning January 4, 2010 and ending May 3, 2020. As expected, the difference in population size causes the coefficients for Swedish deaths to fall below 1. As indicated by the autoregressive parameter at 51 weeks, Norwegian deaths exhibit seasonality over and above those in Sweden. The autoregressive parameters at 1 and 2 weeks suggest a stronger “memory” of high or low values of death in Norway than in Sweden.

**Table 1.** Estimated coefficients and 95% confidence intervals (2-tailed test) for transfer functions predicting all deaths in Norway for 535 weeks beginning January 4, 2010 and ending May 3, 2020.

|  | Estimate | (95% C. I.) |
| --- | --- | --- |
| Constant | 242.34 | 180.34, 304.34 |
| Weekly Swedish Deaths | 0.32 | 0.28, 0.36 |
| Week ending March 22 | -36.85 | -106.85, 33.15 |
| March 29 | -36.47 | -107.59, 34.65 |
| April 5 | -120.94 | -195.14, -46.74 |
| April 12 | -107.15 | -181.25, -33.05 |
| April 19 | -100.52 | -174.02, -27.02 |
| April 26 | -35.45 | -108.85, 37.95 |
| May 3 | -86.39 | -159.67, -13.11 |
| Box Jenkins Parameters |  |  |
| Autoregression at t-1 | 0.15 | 0.06, 0.24 |
| Autoregression at t-2 | 0.21 | 0.12, 0.30 |
| Moving Average at t-51 | 0.14 | 0.05, 0.23 |

Deaths in the second, third, fourth, and sixth weeks following social distancing fell significantly (<u>p</u> < .05, 2-tailed test) below expected. Summing the point estimates of differences suggests that about 414 fewer Norwegians died than expected from deaths in Sweden and from autocorrelation peculiar to Norway in those 4 weeks. Figure 1 shows the differences between observed and expected (i.e., observed – expected) values of Norwegian non-COVID-19 deaths in the last 50 weeks of the test period.

**Figure 1.**
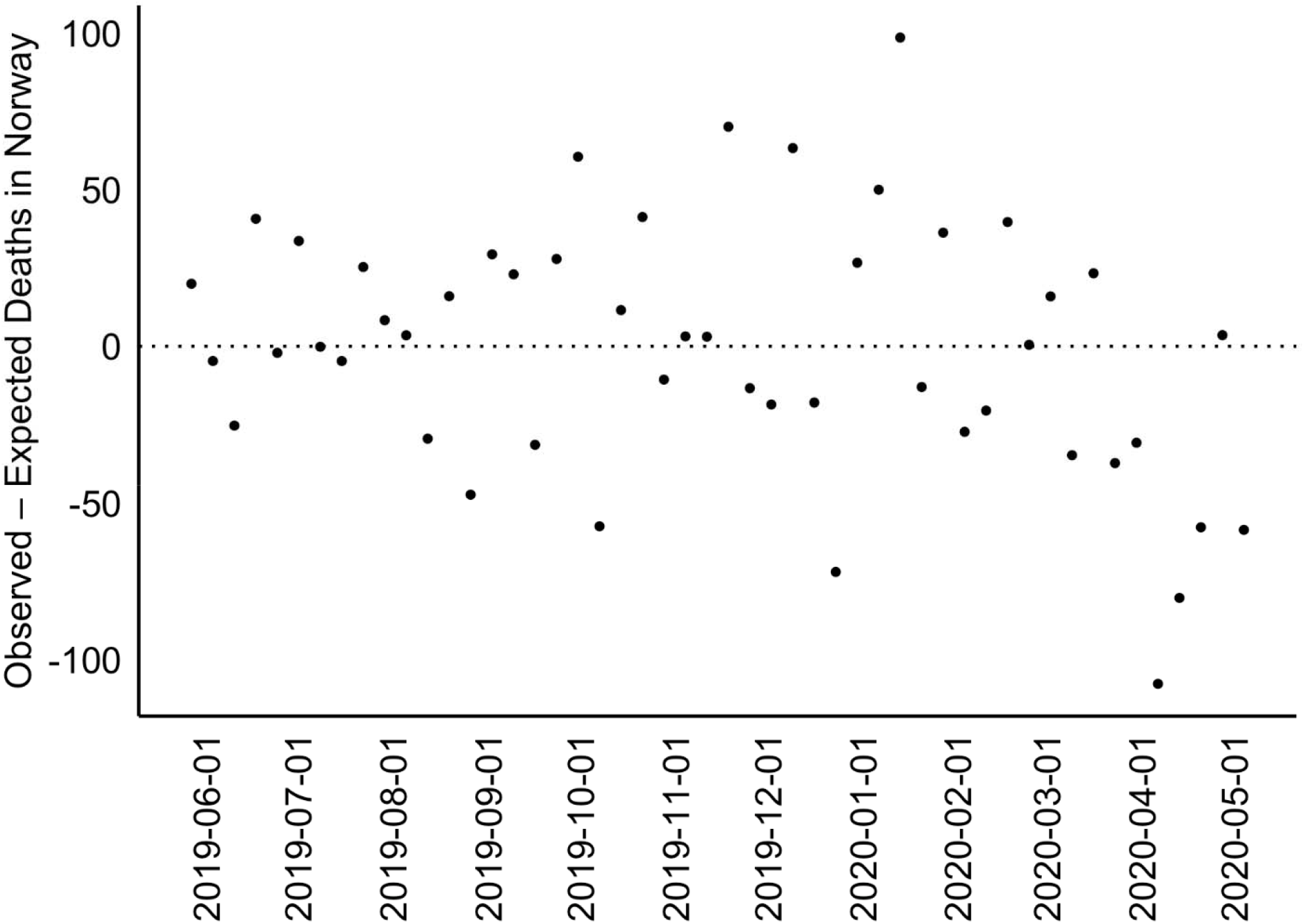
Observed less expected (from Swedish Deaths and autocorrelation) Norwegian non-Covid-19 deaths for 50 weeks ending May 3, 2020 (most recent available data).

To estimate the sensitivity of our analyses to our specification of autocorrelation in deaths, we repeated the steps above but with a “second best fit” Box-Jenkins model used in step 3. The alternative model used the first differences of the Norwegian and Swedish deaths. A moving average parameter at t-1 better fit this specification than did autoregression at t-1 and 1-2 as in the best fitting model. Seasonality expressed with an autoregressive parameter at t-51 remained as in the original model. The results of the test remained essentially same in that deaths in the same weeks as in our primary test appeared significantly lower than expected. Averted deaths summed to 420.

## DISCUSSION

We find that approximately 414 fewer Norwegians than expected died from the hazards of normal life in the 6 weeks following the imposition of social distancing. We derived expected deaths from those in Sweden and from patterns in the history of death in Norway. We do not have cause of death data for the test period so we cannot test speculation as to which hazards declined. Despite liberal retirement benefits beginning at age 62, some Norwegians continue to work until age 67 [8] making the avoidance of work a possible antecedent of averted death. Accidents other than those at work, particularly involving motor vehicles, also likely declined. We cannot, however, rule out that iatrogenic deaths also declined. In Norway, as elsewhere, elective or non-essential use of hospital services fell during the COVID-19 epidemic [9]. An estimated third of hospital deaths in Norway arise from iatrogenic causes [10]. Such deaths likely decreased in the month following the imposition of social distancing.

Strengths of our study include the consistency of methods used to report all-cause mortality in these two Scandinavian countries over a long period. The divergence of policies at a discreet time between these adjacent countries with otherwise similar economic and social policies, further “borrows strength” in estimating lives saved in Norway by social distancing. This circumstance permits the use of rigorous time-series methods to derive counterfactual (i.e., expected) values of weekly mortality in Norway had the government not imposed social distancing. The similarity of results across estimations based on two different models of autocorrelation in counts of weekly deaths further supports the robustness of our findings.

Although not compelled to distance themselves by law, Swedes likely did much to reduce their chances of infection. Using Sweden to generate counterfactual (i.e., expected) values for Norway could, therefore, have biased our estimates towards the null. We may have, as a result, underestimated the true effect of social distancing policies on non-COVID-19-related deaths in Norway.

We focused on Norway and Sweden to gain internal validity for our test. We have, however, no empirical estimate of the external validity of our findings. Further research should estimate the effects of other nations’ policies in response to COVID-19 on all-cause mortality in the short and long term. Delayed care-seeking in response to perceived COVID-19 risk may, for example, decrease iatrogenic deaths in the short run, but increase mortality over time. Based on estimates of seasonality in COVID-19 infections and deaths, such analyses will likely require collection of data well into 2021. More in-depth estimates of the “net effect” of these national measures on other population health indicators (e.g., disability adjusted life years), should prove useful in the debate over the efficacy and prudence of various COVID-19 public health policies.

## Data Availability

All data from public, aggregated, vital statistics sources accessible online

https://www.mortality.org/

https://ourworldindata.org/coronavirus-source-data

## Notes

### Competing Interest Statement

The authors have declared no competing interest.

### Funding Statement

No Funding

